# Recurrence After Hepatic Hydatid Cyst Surgery: Scolicidal Agent Application Technique and the Effect of Cystopiliary Fistula

**DOI:** 10.64898/2026.06.15.26355659

**Authors:** Doğan Erdoğan, Deniz Kol Özbay, Mehmet Ali Özbay

**Affiliations:** Department of General Surgery, Haydarpaşa Numune Training and Research Hospital, University of Health Sciences, Istanbul 34668, Turkey; Department of Surgery, Çay State Hospital, Afyonkarahisar 03700, Turkey; Department of Surgery, Dr. Halil İbrahim Özsoy Bolvadin State Hospital, Afyonkarahisar 03300, Turkey

**Keywords:** Hydatid cyst, Scoliotic agent, Biliary fistula, Recurrence

## Abstract

**Objective:** This study aimed to evaluate long-term outcomes in patients who underwent surgical treatment for hepatic hydatid cyst (HCC) disease and, in particular, to investigate the effect of scolicidal agent (SA) application method and the presence of cystobiliary fistula (CBF) on the development of recurrence.

**Materials and Methods:** This single-center, retrospective study included 197 patients who underwent surgical treatment for HCC disease. Hypertonic saline was used as SA in all patients and was classified as intracystic or pericystic application according to the application method. The presence of CBF was evaluated according to intraoperative and postoperative findings. Patients were followed for 86 months, and the development of recurrence was identified by radiological methods. Comparisons were made between the groups with and without recurrence in terms of SA application method and the presence of CBF.

**Results:** The median age of the patients was 38 years, and the median follow-up period was 86 months. SA application was performed into the cyst in 51.3% of the patients and around the cyst in 48.7%. The presence of CBF was detected in 49.7% of the patients. No statistically significant difference was found between the recurrent and non-recurrent groups in terms of SA application method (p = 0.344). Similarly, no significant relationship was found between the presence of CBF and the development of recurrence (p = 0.721).

**Conclusion:** This study showed that the SA application method and the presence of CBF are not determinants of recurrence in HCC disease. It is thought that recurrence rates can be kept low with appropriate surgical technique and effective biliary tract management.

## 1. Introduction

Hepatic hydatid cyst (HCC) is a significant zoonotic disease caused by Echinococcus granulosus, endemic particularly in livestock-producing regions [1,2]. It remains a serious public health problem in the Mediterranean basin, the Middle East, South America, Africa, and parts of Asia [3]. This disease, which can affect all organs and systems in humans, most frequently affects the liver and lungs [4]. Liver involvement accounts for approximately 60–70% of all cases, and surgical treatment is frequently required in this patient group [5].

The treatment plan includes medical treatment, percutaneous interventions, and surgical approaches depending on the stage, localization, size, and presence of complications of the cyst [6]. Surgical treatment remains the primary treatment option, especially for large and complicated liver cysts [7]. However, postoperative complications and recurrence remain a significant problem despite surgical treatment. In the literature, recurrence rates vary between 2% and 25%, and this wide range is associated with different surgical techniques, patient populations, and follow-up periods [7,8].

Factors that may affect the development of recurrence include the stage of the cyst, the presence of a cystobiliary fistula (CBF), the spread of cyst contents into the peritoneal cavity during surgery, and the scolicidal agents (SA) used [9,10]. In particular, the type and method of application of SA’s play a critical role in the inactivation of protoscoleces. Different SA’s such as hypertonic saline, povidone-iodine, formalin, and silver nitrate have been described in the literature; however, there is no clear consensus on the effectiveness and safety of these agents [10].

One of the most common complications after HCC surgery is CBF, which is associated with bile fistula, infection, and prolonged hospital stay [11,12]. In the literature, there are studies that have identified CBF as a risk factor for late recurrence, as well as studies in which this relationship is not statistically significant [13]. This situation shows that the effect of effectively managing cystobiliary communication surgically on recurrence is still controversial. The aim of this study is to evaluate the long-term follow-up results in patients who underwent surgical treatment for hepatic hydatid cyst disease and to investigate the effect of the scolicidal agent application method using hypertonic saline and the presence of cystobiliary fistula on the development of recurrence. In addition, it was aimed to compare the complications and recurrence rates related to surgical treatment with the literature data.

## 2. Materials and Methods

### 2.1 Study Design and Patient Selection

This study is a single-center, observational study retrospectively examining the data of patients who underwent surgical treatment for HCC in the general surgery clinic of Haydarpaşa Numune Training and Research Hospital. Patients who were operated on for HCC between March 2011 and April 2022 and for whom regular postoperative follow-up data were available were included in the study. Cases of extrahepatic hydatid cysts were included in the study only if they were accompanied by liver involvement.

Diagnosis was made using clinical findings, serological tests, and radiological imaging methods (ultrasonography, computed tomography and/or magnetic resonance imaging). The World Health Organization (WHO) classification was used for staging the cysts [14].

### 2.2 Ethical Approval

This study was conducted in accordance with the principles of the Helsinki Declaration and received ethical approval from the Haydarpaşa Numune Training and Research Hospital Ethics Committee with approval number HNEAH-KAEK/KK/2024/17. Informed patient consent was not required due to the retrospective design of the study.

### 2.3 Surgical Technique and Scoliosis Agent Application

All patients underwent a standard surgical approach performed by the same surgical team, and the surgical technique was planned according to the localization, size, and presence of complications of the cyst. The surgical methods applied included cystotomy-omentoplasty, cystotomy-omentoplasty with cholecystectomy, cysticostomy, or T-tube drainage. Cystectomy and additional surgical procedures were performed in selected cases.

Hypertonic saline solution was used as SA in all cases in the study. SA application was performed either into or around the cyst, according to the surgeon’s preference. There was no heterogeneity among patient groups in terms of SA application method. This provides a more objective evaluation in assessing the effectiveness of SA’s and their effect on recurrence.

### 2.4 Cystobiliary Fistula and Bile Duct Management

The presence of CBF was evaluated considering intraoperative findings and the postoperative clinical course. In patients with detected cystobiliary fistula, bile duct repair was performed in cases where deemed necessary; Preoperative or postoperative endoscopic retrograde cholangiopancreatography (ERCP) was performed in selected patients. Patients requiring bile duct repair and those undergoing additional surgical procedures were recorded.

### 2.5 Postoperative Follow-up and Recurrence Definition

All patients were regularly followed up in the postoperative period. The follow-up period was recorded in months. Recurrence was defined as the detection of a new cyst lesion in the same or a different location during follow-up using radiological imaging methods in patients who had previously undergone surgical treatment. The time to recurrence was recorded in cases where recurrence was detected.

### 2.6 Statistical Analysis

Statistical analyses were performed using SPSS 27.0 (IBM Corp., Armonk, NY, USA). The distribution of continuous variables was assessed using Kolmogorov–Smirnov and Shapiro–Wilk tests. Data showing a normal distribution were expressed as mean ± standard deviation, while data not showing a normal distribution were expressed as median (minimum–maximum). Categorical variables were presented as numbers and percentages.

The relationships between the method of SA administration and the presence of CBF were analyzed using the chi-square test between the relapsed and non-relapsed groups. In all analyses, a p < 0.05 value was considered statistically significant.

## 3. Results

A total of 197 patients who underwent surgical treatment for HCC were included in the study. The median age of the patients was 38 years (12–83), with an average age of 40.5 ± 16.0 years. 50.3% of the patients were female and 49.7% were male. The median follow-up period was 86 months (17–120) (Table 1). When the presenting symptoms of the patients were evaluated, the most common symptom was abdominal pain (79.7%). Nausea-vomiting and jaundice were observed less frequently. In the preoperative period, 9.6% of the patients had a history of recurrence (Table 1). According to the WHO classification, the vast majority of cysts were of the CE2 type (67.0%). The most frequent location was the right hepatic lobe (72.1%). The average largest cyst diameter was 89.1 ± 36.5 mm (Table 1).

**Table 1.** Demographic and Clinical Characteristics of Patients.

| Variable | Min-Max | Median | Mean ± SD / n (%) |
| --- | --- | --- | --- |
| **Age (years)** | 12.0 - 83.0 | 38.0 | 40.5 ± 16.0 |
| **Gender** |  |  |  |
| Female |  |  | 99 (50.3%) |
| Male |  |  | 98 (49.7%) |
| **Follow-up Duration (months)** | 17.0 - 120.0 | 86.0 | 78.3 ± 30.6 |
| **Abdominal Pain** |  |  |  |
| Absent (-) |  |  | 40 (20.3%) |
| Present (+) |  |  | 157 (79.7%) |
| **Nausea and Vomiting** |  |  |  |
| Absent (-) |  |  | 194 (98.5%) |
| Present (+) |  |  | 3 (1.5%) |
| **Jaundice** |  |  |  |
| Absent (-) |  |  | 182 (92.4%) |
| Present (+) |  |  | 15 (7.6%) |
| **History of Pulmonary Hydatid Cyst** |  |  |  |
| Absent (-) |  |  | 194 (98.5%) |
| Present (+) |  |  | 3 (1.5%) |
| **Preoperative Recurrence** |  |  |  |
| Absent (-) |  |  | 178 (90.4%) |
| Present (+) |  |  | 19 (9.6%) |
| **Length of Hospital Stay (days)** | 3.0 - 41.0 | 8.0 | 10.4 ± 7.2 |
| **GGT (U/L)** | 7.0 - 801.0 | 34.0 | 57.1 ± 80.7 |
| **ALP (U/L)** | 34.0 - 404.0 | 81.0 | 96.0 ± 53.3 |
| **Total Bilirubin (mg/dL)** | 0.1 - 6.9 | 0.6 | 0.8 ± 0.7 |
| **Direct Bilirubin (mg/dL)** | 0.1 - 6.7 | 0.3 | 0.4 ± 0.6 |
| **WBC ($\times 10^3/\mu\text{L}$)** | 3110 - 22000 | 7940 | 8515.2 ± 2712.7 |
| **WHO Classification** |  |  |  |
| CE2 |  |  | 132 (67.0%) |
| CE3A |  |  | 27 (13.7%) |
| CE3B |  |  | 17 (8.6%) |
| CE4 |  |  | 15 (7.6%) |
| CE5 |  |  | 6 (3.0%) |
| **Radiological Imaging** |  |  |  |
| **USG** |  |  |  |
| Absent (-) |  |  | 96 (48.7%) |
| Present (+) |  |  | 101 (51.3%) |
| **CT** |  |  |  |
| Absent (-) |  |  | 104 (52.8%) |
| Present (+) |  |  | 93 (47.2%) |
| **MRI / MRCP** |  |  |  |
| Absent (-) |  |  | 120 (60.9%) |
| Present (+) |  |  | 77 (39.1%) |
| **Largest Cyst Diameter (mm)** | 30.0 - 270.0 | 80.0 | 89.1 ± 36.5 |
| **Cyst Location** |  |  |  |
| Right Lobe |  |  | 142 (72.1%) |
| Left Lobe |  |  | 26 (13.2%) |
| Both Lobes |  |  | 29 (14.7%) |

*SD: Standard Deviation; WHO: World Health Organization; USG: Ultrasonography; CT: Computed Tomography MRI/MRCP: Magnetic Resonance Imaging/Magnetic Resonance Cholangiopancreatography.*

The presence of extrahepatic cysts was detected in 12.7% of cases, with the most frequent extrahepatic involvement being the spleen (9.1%). Preoperative ERCP was performed in 13.2% of cases, and postoperative ERCP in 6.1% (Table 2). When the surgical methods applied were examined, cystotomy + omentoplasty was performed in 66.0% of patients, and cystotomy + omentoplasty + cholecystectomy in 29.4%. The cystectomy rate was 8.6%. Bile duct repair was performed in 34.5% of patients (Table 2).

**Table 2.**
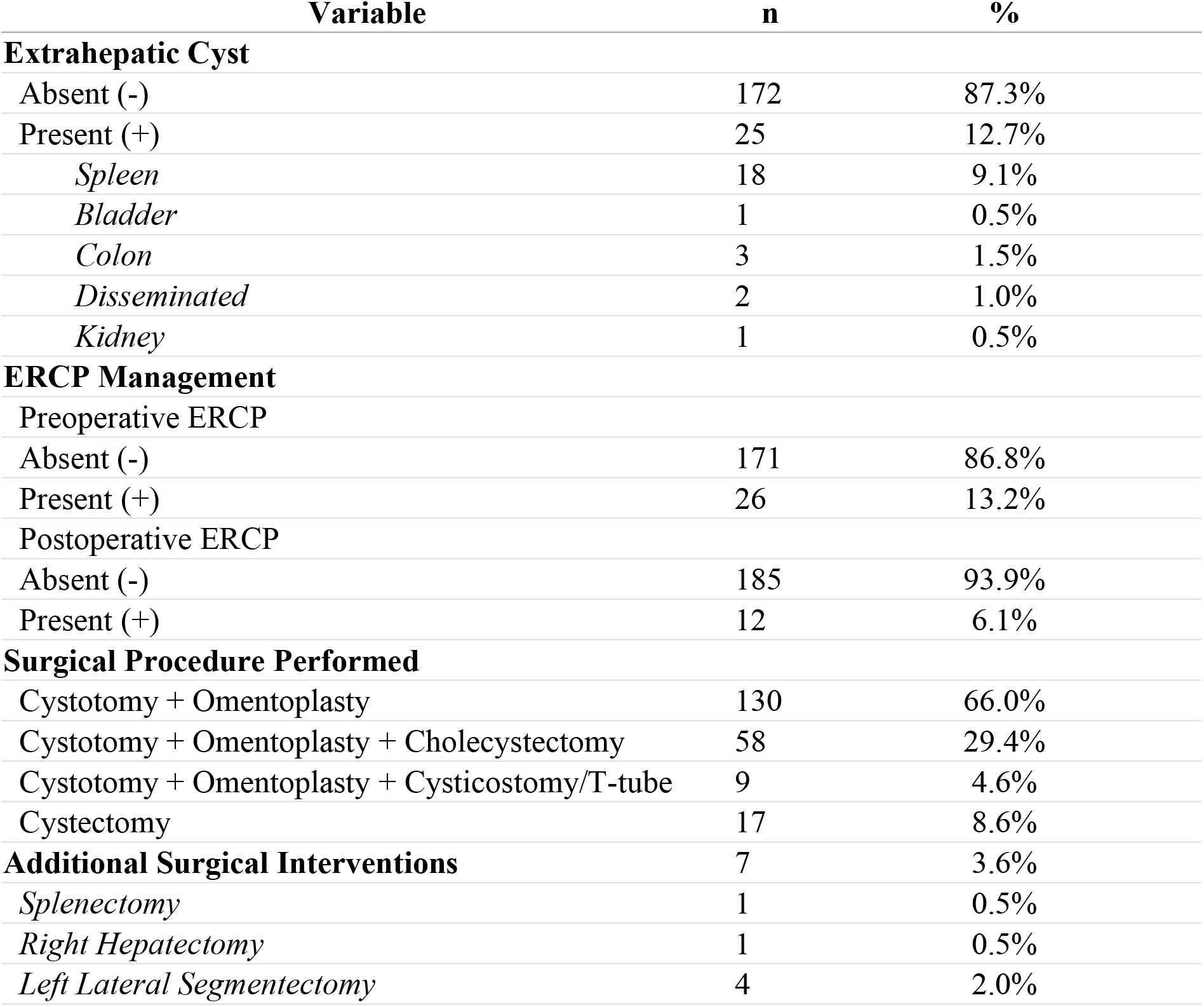
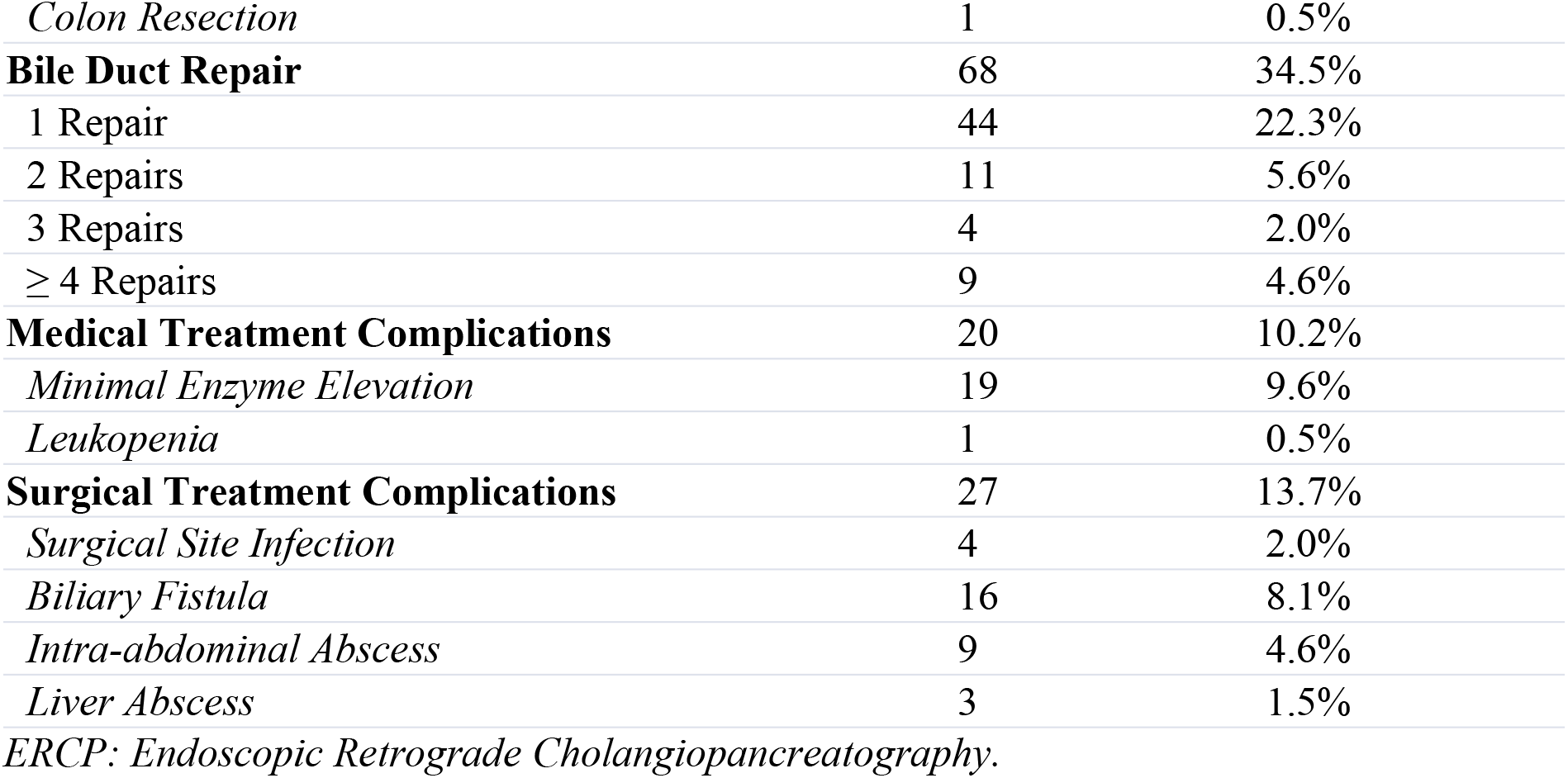
Operative and Clinical Characteristics.

Surgery-related complications were detected in 13.7% of cases. The most frequent postoperative complication was bile fistula (8.1%). The peroperative complication rate was recorded as 6.1% (Table 3). SA application was performed in all patients using hypertonic saline, applied into the cyst in 51.3% and around the cyst in 48.7%. CBF presence was detected in 49.7% of patients (Table 3).

**Table 3.** Perioperative Findings and Recurrence Parameters.

| Variable | Min-Max | Median | Mean ± SD / n (%) |
| --- | --- | --- | --- |
| **Perioperative Complications** |  |  |  |
| Present (+) |  |  | 12 (6.1%) |
| *Duodenal Injury* |  |  | 1 (0.5%) |
| *Diaphragmatic Injury* |  |  | 10 (5.1%) |
| *Colon Injury* |  |  | 1 (0.5%) |
| **Time to Recurrence (months)** | 7.0 - 72.0 | 31.0 | 38.0 ± 25.6 |
| **Scolocidal Agent Usage** |  |  |  |
| Intracystic Application |  |  | 101 (51.3%) |
| Pericystic Application |  |  | 96 (48.7%) |
| **Cystobiliary Fistula** |  |  |  |
| Absent (-) |  |  | 99 (50.3%) |
| Present (+) |  |  | 98 (49.7%) |

When recurrence development was evaluated, no statistically significant difference was found between the groups with and without recurrence in terms of SA application method (p = 0.344). Similarly, no significant relationship was found between CBF and recurrence development (p = 0.721) (Table 4).

**Table 4.** Comparison of Factors Associated with Recurrence.

| Variable | Recurrence (-) (n=188)<br>n (%) | Recurrence (+) (n=9)<br>n (%) | p-value |
| --- | --- | --- | --- |
| **Scolocidal Agent Usage** |  |  | 0.344 |
| Intracystic Application | 95 (50.5%) | 6 (66.7%) |  |
| Pericystic Application | 93 (49.5%) | 3 (33.3%) |  |
| **Cystobiliary Fistula** |  |  | 0.721 |
| Absent (-) | 95 (50.5%) | 4 (44.4%) |  |
| Present (+) | 93 (49.5%) | 5 (55.6%) |  |

*Statistical analysis performed using Chi-square ( $\chi^2$ ) test. A p-value > 0.05 indicates no significant difference between groups.*

## 4. Discussion

Our study was conducted retrospectively in a large patient group and includes long-term results of patients who underwent surgical treatment for HCC disease. In particular, the effect of the method of scolicidal agent application and the presence of cystobiliary fistula on the development of recurrence was investigated.

SA’s used during surgery both provide inactivation of protoscoleces and play a critical role in preventing intraabdominal spread [15]. Different SA’s and application techniques have been described in studies, and variable results have been reported regarding the effectiveness of these agents [16]. Although intracystic injection of scolicidal agents into the cyst has been widely used in past studies, intracystic injection has not been recommended in surgery due to limited objective evidence regarding their effectiveness and potential toxicity [17]. In the current study by Sharafi et al., the effectiveness and application method of several SA’s, primarily hypertonic saline, were investigated, but it was emphasized that a standard approach could not be found [10]. In our study, hypertonic saline, one of the most commonly used agents for SA today, was used in all patients, eliminating heterogeneity depending on the type of agent and allowing for a more objective evaluation of the effect of the application method on recurrence. There is no similar study in the literature evaluating the relationship between application technique and recurrence for SA, and in our study, it was observed that there was no statistically significant difference between application into and around the cyst.

One of the most common complications after HCC surgery is cystobiliary fistula [18]. In patients with cystobiliary fistula, infection and prolonged hospital stays are observed. Kara et al. reported in their study that cystobiliary communication especially increases the risk of postoperative complications [19]. However, when the literature is examined, the relationship between cystobiliary fistula and recurrence is not entirely clear. Although Habeeb et al. reported a relationship between late-stage recurrence and cystobiliary fistula in some cases, this finding has not been confirmed in all series [9]. The significant variability in the literature highlights the importance of thorough preoperative radiological evaluation to identify cystobiliary communication and interventions using methods like ERCP, as well as the application of correct surgical techniques to cystobiliary communication detected intraoperatively, in preventing CBF formation. The lack of a significant correlation between CBF presence and recurrence in our study, despite a long follow-up period, suggests that the risk of recurrence can be reduced by applying appropriate endoscopic interventions and performing effective bile duct repair.

Our study has a large patient series and a long follow-up period, and the recurrence rates we found are consistent with the literature, which is clinically valuable. Standardization of surgical techniques, homogeneous use of biliary tract agents, and effective bile duct management may have contributed to keeping recurrence rates low.

Our study has some limitations. As it is a retrospective study, there is a possibility of data loss. Also, being a single-center study may limit the universality of the results. However, the high number of patients and the presentation of long-term follow-up results are among the strengths of the study.

## 5. Conclusions

In our study, data from patients who underwent surgical treatment for hydatid cyst disease were retrospectively examined, and long-term outcomes and factors that may be associated with recurrence were investigated. According to the results of our study, the method of saline administration using hypertonic saline and the presence of biliary tract complications did not show a statistically significant relationship with the development of recurrence.

These findings suggest that the method of saline administration alone is not a determining factor in recurrence in hydatid cyst surgery, and that the presence of biliary tract complications can be effectively controlled with appropriate surgical and biliary tract management. In light of our data, standardization of surgical techniques, homogenous use of saline, and effective management of biliary tract complications have shown us that they will play an important role in keeping recurrence rates low.

In conclusion, surgical experience, appropriate techniques, and multidisciplinary approaches in hydatid cyst surgery can prevent recurrences after surgical treatment. Multicenter and prospective studies in this field will contribute to a clearer understanding of the factors affecting the risk of recurrence.

## Data Availability

All relevant data underlying the findings of this study are contained within the manuscript and its Supporting Information files. Due to ethical and privacy restrictions related to patient confidentiality, individual-level de-identified data cannot be made publicly available. Qualified researchers may request access to de-identified data from the corresponding author, subject to approval by the relevant Ethics Committee and institutional regulations.

## Author Contributions

Conceptualization, D.E.; methodology, D.K.Ö. and D.E.; data collection, D.E.; data curation, D.K.Ö.; formal analysis, D.K.Ö. and M.A.Ö.; investigation, D.K.Ö.; writing-original draft preparation, D.K.Ö.; writing-review and editing, M.A.Ö.; supervision, D.E.; project administration, D.E. All authors have read and agreed to the published version of the article.

## Funding

This research received no external funding.

## Institutional Review Board Statement

Our study was conducted in accordance with the Helsinki Declaration and received approval from the ethics committee of Haydarpaşa Numune Training and Research Hospital (protocol code HNEAH-KAEK/KK/2024/17 and date: 12.02.2024).

## Informed Consent Statement

This study was a retrospective review of medical records. Patient data were accessed after approval from the Haydarpaşa Numune Training and Research Hospital Ethics Committee (Approval No: HNEAH-KAEK/KK/2024/17). The Ethics Committee waived the requirement for informed consent due to the retrospective nature of the study. The authors had access to identifiable information during data collection; however, all data were anonymized and de-identified prior to analysis and reporting.

## Data Availability Statement

Data obtained during the research process may be shared with researchers who request it, in accordance with ethical and legal frameworks.

## Conflicts of Interest

The authors declare no conflicts of interest.

## Disclaimer/Publisher’s Note

The statements, opinions and data contained in all publications are solely those of the individual author(s) and contributor(s) and not of PLOS and/or the editor(s). PLOS and/or the editor(s) disclaim responsibility for any injury to people or property resulting from any ideas, methods, instructions or products referred to in the content.

## Notes

### Competing Interest Statement

The authors have declared no competing interest.

### Funding Statement

The author(s) received no specific funding for this work.

### Author Declarations

This retrospective study was approved by the Haydarpaşa Numune Training and Research Hospital Clinical Research Ethics Committee (Approval No: HNEAH-KAEK 2024/17, Date: 12.02.2024). The requirement for informed consent was waived by the ethics committee due to the retrospective design of the study and the use of anonymized patient data.

